# Are rare heterozygous *SYNJ1* variants associated with Parkinson’s disease?

**DOI:** 10.1101/2024.05.29.24307986

**Authors:** Konstantin Senkevich, Sitki Cem Parlar, Cloe Chantereault, Eric Yu, Jamil Ahmad, Jennifer A. Ruskey, Farnaz Asayesh, Dan Spiegelman, Cheryl Waters, Oury Monchi, Yves Dauvilliers, Nicolas Dupré, Irina Miliukhina, Alla Timofeeva, Anton Emelyanov, Sofya Pchelina, Lior Greenbaum, Sharon Hassin-Baer, Roy N. Alcalay, Ziv Gan-Or

## Abstract

Previous studies have suggested that rare biallelic *SYNJ1* mutations may cause autosomal recessive parkinsonism and Parkinson’s disease (PD). Our study explored the impact of rare *SYNJ1* variants in non-familial settings, including 8,165 PD cases, 818 early-onset PD (EOPD, <50 years) and 70,363 controls. Burden meta-analysis using optimized sequence Kernel association test (SKAT-O) revealed an association between rare nonsynonymous variants in the Sac1 SYNJ1 domain and PD (P_fdr_=0.040). Additionally, a meta-analysis focusing on patients with EOPD demonstrated an association between all rare *SYNJ1* variants and PD (P_fdr_=0.029). Rare *SYNJ1* variants may be associated with sporadic PD, and more specifically with EOPD.

---

Parkinson’s disease (PD) is a complex condition influenced by multiple genes and environmental factors, with specific single-gene mutations accounting only for 1–2% of cases, excluding *GBA1*^1^. Early onset of PD (EOPD; <50 years) represents about 3-14% of all PD cases depending on population ^2^, and has been linked to mutations in several genes (i.e. *PRKN, PINK1, DJ-1, LRRK2, SNCA, GBA1*) ^2^. Considering variants in these genes only, the frequency of genetically associated PD in early-onset cases reaches about 16.6% ^3^.

Synaptojanin 1, encoded by *SYNJ1*, is a lipid phosphatase abundantly expressed in brain tissues ^4^ which plays an important role in synaptic trafficking and in autophagic clearance ^5-7^. The protein consists of three functionally distinct domains: Sac1, 5-phosphatase, and proline rich domains (PRD) ^8^. Previous studies suggested that biallelic *SYNJ1* mutations cause autosomal recessive, early-onset parkinsonism and EOPD ^9-12^. In the current study, we sought to examine the role of rare *SYNJ1* variants in six non-familial cohorts with a total of 8,165 PD cases (including 818 EOPD) and 70,363 controls (detailed in Supplementary Table 1).

The average coverage of the *SYNJ1* gene in the four cohorts sequenced at McGill was >4000X, with 100% of nucleotides covered at >30X (Supplementary Table 2). From these cohorts, 23-75 rare variants were included in the analysis, depending on the cohort (detailed in Supplementary Table 3). In the AMP-PD cohort, 523 rare variants were included in the analysis and 440 rare variants were included in the UKBB cohort (coding and functional variants detailed in Supplementary Table 4).

Burden analysis with SKAT-O suggested a possible association between all rare variants and PD in the AMP-PD cohort (P=1.42E-05; P_fdr_=2.98E-04), with nominal associations that did not survive false discovery rate (fdr) correction in the Columbia cohort (P=0.030; P_fdr_=0.126) and the McGill cohort (P=0.009; P_fdr_=0.095). No association was found in the meta-analysis of all six cohorts after false discovery rate correction (P=0.025; P_fdr_=0.131). We also found a nominal association between non-synonymous variants and PD in the Columbia cohort (P=0.012; P_fdr_=0.084; Table 1).

**Table 1.**
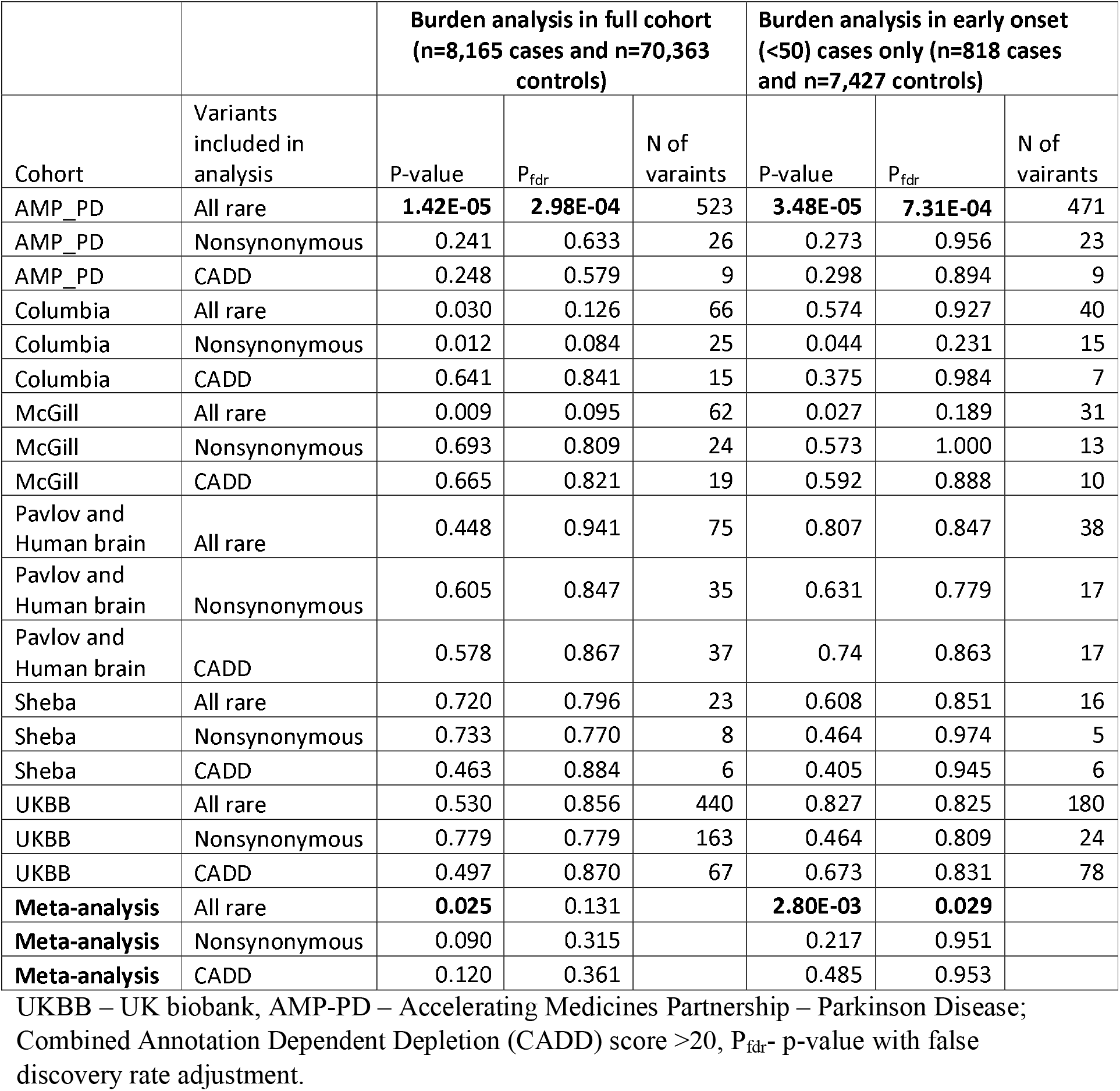
Burden analysis of rare *SYNJ1* variants

We then analyzed only EOPD (age at onset < 50 years) and found an association between all rare variants and PD in the AMP-PD cohort (P=3.48E-05; P_fdr_=7.31E-04) and nominal association that did not survive FDR correction in the McGill cohort (P=0.027; P_fdr_=0.189). In the meta-analysis, which included only EOPD patients and controls, all rare variants in *SYNJ1* were associated with PD (P=2.80E-03; P_fdr_=0.029). The association between non-synonymous variants and PD was nominally significant in the Columbia cohort (P=0.044; P_fdr_=0.231) but not in the meta-analysis (Table 1).

To analyze variants within specific functional domains of SYNJ1 (Sac1, 5-phosphatase, PRD), we divided the gene regions into these domains and then repeated SKAT-O analysis. We found an association between the Sac1 domain of SYNJ1 and PD for non-synonymous variants with high CADD scores in the UKBB cohort (P=0.006; P_fdr_=0.082), which did not survive fdr correction. However, this association became significant in the meta-analysis (P=0.002; P_fdr_=0.04; Supplementary Table 5). We observed that this association was primary driven by the p.A195T *SYNJ1* variant in the UKBB cohort (Odds ratio (OR)=4.87; 95% confidence interval (CI) 1.76-13.46, P=0.002, with a minor allele frequency (MAF) in cases of 0.001 and 0.0002 in controls and gnomAD). This variant is classified as a Variant of Uncertain Significance (VUS) in ClinVar and is likely benign according to the ACMG classification but is noted for its high CADD score. This variant was also detected in one patient from the Pavlov and Human Brain cohort and was not reported in any other studied cohorts.

We did not find any homozygous or compound heterozygous carriers of pathogenic variants (previously reported in association with PD ^10^, defined as pathogenic or likely pathogenic by ClinVar or variants with CADD score >20) in none of the studied cohorts.

In the current study, we found an association between all rare heterozygous variants in *SYNJ1* and heterozygous variants with high CADD score in the Sac1 domain of SYNJ1, and the risk of PD in some of the analyzed cohorts. These findings suggest that *SYNJ1* could be associated with sporadic PD. Additional studies are required to determine whether this potential association holds in other cohorts. The association we found of rare heterozygous *SYNJ1* variants with EOPD was more convincing, yet here too, additional studies are needed. We did not identify biallelic carriers in our analysis, although private bi-allelic *SYNJ1* variants were previously associated with EOPD and atypical parkinsonism (detailed in ^10^),

Multiple genes that are involved in the autophagy-lysosomal pathway are also associated with PD ^13^. From the biological point of view, *SYNJ1* has a role in two pathways relevant to PD: synaptic trafficking and autophagic clearance ^6,7^. Recent functional studies demonstrated that mutations in *SYNJ1* destabilize dopaminergic neurons potentially due to defective clathrin uncoating, disrupting lipid metabolism and synaptic function ^14,15^. However, SYNJ1 overexpression can counteract these effects, highlighting its potential therapeutic potential in PD^14,15^.

Our study has several limitations. In some of our cohorts, we had significant differences in sex and age between PD patients and controls. We adjusted for the effects of these covariates in our analysis to address this limitation. Additionally, in our study we predominantly included participants with European ancestry. Last, we different types of sequencing data and quality control procedures were performed independently for each of the cohorts. Thus, it could potentially create differences in variant enrichment between cohorts. To partially overcome this limitation, we analyzed each cohort separately and conducted meta-analyses of the different cohorts, rather than joint analysis of all cohorts.

To conclude, we found that rare heterozygous *SYNJ1* variants were potentially associated with EOPD and variants in the Sac1 domain are associated with sporadic PD. Larger studies in cohorts of different ethnic backgrounds are needed to replicate our results.

## Methods

### Population

The study population comprised 8,165 PD patients and 70,363 controls from six cohorts, including 818 EOPD (all demographic characteristics detailed in Supplementary Table 1). Four cohorts were collected at McGill university: (1) a cohort of French/French-Canadian from Quebec, Canada and Montpellier, France, (2) a cohort from Columbia University (New York, NY), (3) a cohort from Sheba Medical Center (Israel) and (4) a cohort from Pavlov and Human Brain institutes (Russia). ^16^The Columbia cohort comprises patients and controls of varied racial and ethnic origin (European, Ashkenazi [AJ] descent and a minority of Hispanics and blacks). Patients and controls in the Sheba cohort, which was recruited in Israel, are of full AJ ancestry (all four grandparents are AJ). The Pavlov and Human Brain institute cohort was collected from the North-Western region of Russia and mainly with East-European ancestry. Additionally, we performed the analysis in the Accelerating Medicines Partnership – Parkinson Disease (AMP-PD) initiative cohorts (https://amp-pd.org/; detailed in the Acknowledgment) and the UK biobank (UKBB) cohort. We only included participants of European ancestry from both cohorts, and we excluded any first and second-degree relatives from the analysis.

All PD patients were diagnosed by movement disorder specialists according to the UK brain bank criteria ^17^ or the MDS clinical diagnostic criteria ^18^.

### Standard Protocol Approvals, Registrations, and Patient Consents

All local IRBs approved the protocols and informed consent was obtained from all individual participants before entering the study.

### Targeted next-generation sequencing by molecular inversion probes

The entire coding sequence of the *SYNJ1* gene, including exon-intron boundaries (±50bps) and the 5’ and 3’ untranslated regions (UTRs), was targeted using molecular inversion probes (MIPs) as described earlier ^19^. The full protocol is available at https://github.com/gan-orlab/MIP_protocol. The Genome Quebec Innovation Centre’s Illumina NovaSeq 6000 SP PE100 platform was used to sequence the library. Alignment was carried out to hg19 reference genome ^20^ with coordinates for *SYNJ1* chr21:34,001,069-34,100,351. Genome Analysis Toolkit (GATK, v3.8) was used for post-alignment quality checking and variant calling ^21^. We applied standard quality control procedures as described before ^22^. In brief, using the PLINK program version 1.9 and GATK, v3.8, we carried out quality control by eliminating variants and samples with poor quality. SNPs and samples with genotyping rate lower than 90% were excluded. The analyses only included variants with 30x minimal depths of coverage, having a MAF less than 1% and a minimum quality score (GQ) of 30.

### Whole-exome and whole-genome sequencing data

The genetic data in AMP-PD and UKBB were aligned to the human reference genome hg38 and we used the appropriate coordinates to extract the *SYNJ1* data (chr21:32,628,759-32,727,939). Quality control procedures were performed as previously described in detail for AMP-PD ^23^ and UKBB ^24^ cohorts.

### Statistical Analysis

To analyze rare variants (MAF < 0.01), we applied the optimized sequence Kernel association test (SKAT-O, R package) ^25^ in each cohort separately, followed by meta-analysis using the metaSKAT package ^26^. We performed separate analyses for all rare variants, non-synonymous variants, and variants with Combined Annotation Dependent Depletion (CADD) scores of ≥ 20. For domain-based analysis, domains boundaries were decided by the widest intervals of each domain based on a combination of estimates from publicly available domain annotation resources. These resources are SuperFamily, Pfam, Smart, Gene3D, PANTHER, Conserved Domains Database, and PROSITE ^27-30^. We adjusted for sex, age and ethnicity in all analyses. FDR correction was applied to all p-values.

## Data Availability

All data relevant to this study are included within thearticle and the accompanying supplementary files.

## Data availability

Data used in the preparation of this article were obtained from the AMP PD Knowledge Platform (https://www.amp-pd.org) and UKBB. Access to these datasets is available for eligible researchers upon request. The code utilized in this study is accessible at https://github.com/gan-orlab/SYNJ1. We provided the variants used for burden analyses in the supplementary tables.

## Acknowledgements

We would like to thank the participants in the different cohorts for contributing to this study. This research used the NeuroHub infrastructure and was undertaken thanks in part to funding from the Canada First Research Excellence Fund, awarded through the Healthy Brains, Healthy Lives initiative at McGill University, Calcul Québec and Compute Canada. This research has been conducted using the UK Biobank Resource under Application Number 45551. Data used in the preparation of this article were obtained from the Accelerating Medicine Partnership® (AMP®) Parkinson’s Disease (AMP PD) Knowledge Platform. For up-to-date information on the study, visit https://www.amp-pd.org. The AMP® PD program is a public-private partnership managed by the Foundation for the National Institutes of Health and funded by the National Institute of Neurological Disorders and Stroke (NINDS) in partnership with the Aligning Science Across Parkinson’s (ASAP) initiative; Celgene Corporation, a subsidiary of Bristol-Myers Squibb Company; GlaxoSmithKline plc (GSK); The Michael J. Fox Foundation for Parkinson’s Research ; Pfizer Inc.; Sanofi US Services Inc.; Verily Life Sciences and AbbVie. ACCELERATING MEDICINES PARTNERSHIP and AMP are registered service marks of the U.S. Department of Health and Human Services. Genetic data used in preparation of this article were obtained from the Fox Investigation for New Discovery of Biomarkers (BioFIND), the Harvard Biomarker Study (HBS), the Parkinson’s Progression Markers Initiative (PPMI), the Parkinson’s Disease Biomarkers Program (PDBP), the International LBD Genomics Consortium (iLBDGC), and the STEADY-PD III Investigators. BioFIND is sponsored by The Michael J. Fox Foundation for Parkinson’s Research (MJFF) with support from the National Institute for Neurological Disorders and Stroke (NINDS). The BioFIND Investigators have not participated in reviewing the data analysis or content of the manuscript. For up-to-date information on the study, visit michaeljfox.org/news/biofind. The Harvard Biomarker Study (HBS) is a collaboration of HBS investigators [full list of HBS investigators found at https://www.bwhparkinsoncenter.org/biobank/ and funded through philanthropy and NIH and Non-NIH funding sources. The HBS Investigators have not participated in reviewing the data analysis or content of the manuscript. PPMI is sponsored by The Michael J. Fox Foundation for Parkinson’s Research and supported by a consortium of scientific partners: [list the full names of all of the PPMI funding partners found at https://www.ppmi-info.org/about-ppmi/who-we-are/study-sponsors]. The PPMI investigators have not participated in reviewing the data analysis or content of the manuscript. For up-to-date information on the study, visit www.ppmi-info.org. The Parkinson’s Disease Biomarker Program (PDBP) consortium is supported by the National Institute of Neurological Disorders and Stroke (NINDS) at the National Institutes of Health. A full list of PDBP investigators can be found at https://pdbp.ninds.nih.gov/policy. The PDBP investigators have not participated in reviewing the data analysis or content of the manuscript. The Study of Isradipine as a Disease-modifying Agent in Subjects With Early Parkinson Disease, Phase 3 (STEADY-PD3) is funded by the National Institute of Neurological Disorders and Stroke (NINDS) at the National Institutes of Health with support from The Michael J. Fox Foundation and the Parkinson Study Group. For additional study information, visit https://clinicaltrials.gov/ct2/show/study/NCT02168842. The STEADY-PD3 investigators have not participated in reviewing the data analysis or content of the manuscript. Genome sequence data for the Lewy body dementia case-control cohort were generated at the Intramural Research Program of the U.S. National Institutes of Health. The study was supported in part by the National Institute on Aging (program #: 1ZIAAG000935) and the National Institute of Neurological Disorders and Stroke (program #: 1ZIANS003154). ZGO is supported by the Fonds de recherche du Québec - Santé (FRQS) Chercheurs-boursiers award, and is a William Dawson Scholar. The access to part of the participants for this research has been made possible thanks to the Quebec Parkinson’s Network (http://rpq-qpn.ca/en/). KS is supported by Parkinson Canada Movement Disorders clinical fellowship.

## Author contributions

KS was responsible for the design and conceptualization of the study, acquisition, and analysis of data, and drafting or revising the manuscript for intellectual content. ZGO was responsible for the design and conceptualization of the study, acquisition, and analysis of data, and drafting or revising the manuscript for intellectual content. SCP, CC, EY, JA, JAR, FA, DS, CW, OM, YD, ND, IM, AT, AE, SP, LG, SHB, and RNA were involved in the acquisition and analysis of data and in drafting or revising the manuscript for intellectual content. All authors read and approved the final manuscript.

## References

1 Blauwendraat, C., Nalls, M. A. & Singleton, A. B. The genetic architecture of Parkinson’s disease. The Lancet Neurology 19, 170–178 (2020).

2 Riboldi, G. M., Frattini, E., Monfrini, E., Frucht, S. J. & Di Fonzo, A. A Practical Approach to Early-Onset Parkinsonism. Journal of Parkinson’s Disease 12, 1–26 (2022). 10.3233/JPD-212815

3 Alcalay, R. N. et al. Frequency of Known Mutations in Early-Onset Parkinson Disease: Implication for Genetic Counseling: The Consortium on Risk for Early Onset Parkinson Disease Study. Archives of Neurology 67, 1116–1122 (2010). 10.1001/archneurol.2010.194

4 Miranda, A. M. et al. Excess synaptojanin 1 contributes to place cell dysfunction and memory deficits in the aging hippocampus in three types of Alzheimer’s disease. Cell reports 23, 2967–2975 (2018).

5 Fasano, D. et al. Alteration of endosomal trafficking is associated with early-onset parkinsonism caused by SYNJ1 mutations. Cell Death & Disease 9, 385 (2018). 10.1038/s41419-018-0410-7

6 Choudhry, H., Aggarwal, M. & Pan, P.-Y. Mini-review: Synaptojanin 1 and its implications in membrane trafficking. Neuroscience Letters 765, 136288 (2021). 10.1016/j.neulet.2021.136288

7 George, A. A. et al. Synaptojanin 1 is required for endolysosomal trafficking of synaptic proteins in cone photoreceptor inner segments. PloS one 9, e84394 (2014).

8 Chang-Ileto, B. & Di Paolo, G. in Encyclopedia of Neuroscience (ed Larry R. Squire) 809–814 (Academic Press, 2009).

9 Olgiati, S. et al. PARK20 caused by SYNJ1 homozygous Arg258Gln mutation in a new Italian family. neurogenetics 15, 183–188 (2014).

10 Lesage, S. et al. Clinical Variability of SYNJ1-Associated Early-Onset Parkinsonism. Frontiers in neurology 12, 648457 (2021).

11 Xie, F. et al. A novel homozygous SYNJ1 mutation in two siblings with typical Parkinson’s disease. Parkinsonism & Related Disorders 69, 134–137 (2019).

12 Kumar, S. et al. Novel and reported variants in Parkinson’s disease genes confer high disease burden among Indians. Parkinsonism & Related Disorders 78, 46–52 (2020).

13 Senkevich, K. & Gan-Or, Z. Autophagy lysosomal pathway dysfunction in Parkinson’s disease; evidence from human genetics. Parkinsonism & related disorders 73, 60–71 (2020).

14 Jacquemyn, J. et al. Parkinsonism mutations in DNAJC6 cause lipid defects and neurodegeneration that are rescued by Synj1. npj Parkinson’s Disease 9, 19 (2023). 10.1038/s41531-023-00459-3

15 Ng, X. Y. et al. Mutations in Parkinsonism-linked endocytic proteins synaptojanin1 and auxilin have synergistic effects on dopaminergic axonal pathology. npj Parkinson’s Disease 9, 26 (2023). 10.1038/s41531-023-00465-5

16 Gan-Or, Z. et al. The Quebec Parkinson Network: A Researcher-Patient Matching Platform and Multimodal Biorepository. J Parkinsons Dis 10, 301–313 (2020). 10.3233/jpd-191775

17 Hughes, A. J., Ben-Shlomo, Y., Daniel, S. E. & Lees, A. J. What features improve the accuracy of clinical diagnosis in Parkinson’s disease. A clinicopathologic study 42, 1142–1142 (1992). 10.1212/wnl.42.6.1142

18 Postuma, R. B. et al. MDS clinical diagnostic criteria for Parkinson’s disease. Movement disorders 30, 1591–1601 (2015).

19 Rudakou, U. et al. Targeted sequencing of Parkinson’s disease loci genes highlights SYT11, FGF20 and other associations. Brain 144, 462–472 (2021). 10.1093/brain/awaa401

20 Li, H. & Durbin, R. Fast and accurate short read alignment with Burrows–Wheeler transform. Bioinformatics 25, 1754–1760 (2009). 10.1093/bioinformatics/btp324

21 McKenna, A. et al. The Genome Analysis Toolkit: a MapReduce framework for analyzing next-generation DNA sequencing data. Genome research 20, 1297–1303 (2010).

22 Bencheikh, B. O. A. et al. Variants in the Niemann-Pick type C gene NPC1 are not associated with Parkinson’s disease. Neurobiology of Aging (2020).

23 Iwaki, H. et al. Accelerating medicines partnership: Parkinson’s disease. genetic resource. Movement Disorders 36, 1795–1804 (2021).

24 Carson, A. R. et al. Effective filtering strategies to improve data quality from population-based whole exome sequencing studies. BMC Bioinformatics 15, 125 (2014). 10.1186/1471-2105-15-125

25 Lee, S. et al. Optimal unified approach for rare-variant association testing with application to small-sample case-control whole-exome sequencing studies. Am J Hum Genet 91, 224–237 (2012). 10.1016/j.ajhg.2012.06.007

26 Lee, S., Teslovich, T. M., Boehnke, M. & Lin, X. General framework for meta-analysis of rare variants in sequencing association studies. Am J Hum Genet 93, 42–53 (2013). 10.1016/j.ajhg.2013.05.010

27 Pandurangan, A. P., Stahlhacke, J., Oates, M. E., Smithers, B. & Gough, J. The SUPERFAMILY 2.0 database: a significant proteome update and a new webserver. Nucleic acids research 47, D490–D494 (2019).

28 Mi, H. et al. PANTHER version 11: expanded annotation data from Gene Ontology and Reactome pathways, and data analysis tool enhancements. Nucleic acids research 45, D183–D189 (2017).

29 Marchler-Bauer, A. et al. CDD: a Conserved Domain Database for protein classification. Nucleic acids research 33, D192–D196 (2005).

30 Sigrist, C. J. et al. New and continuing developments at PROSITE. Nucleic Acids Res 41, D344–347 (2013). 10.1093/nar/gks1067

